# Potentially Inappropriate Medication Use and the Hospitalization Rate among Thai Elderly Patients: a Retrospective Cohort Study

**DOI:** 10.1101/2021.09.13.21263490

**Authors:** Vorawee Varavithya, Chayanee Tirapat, Penpitcha Rojpibulstit, Panadda Poovichayasumlit, Vanida Prasert, Pasitpon Vatcharavongvan

**Affiliations:** Faculty of Medicine, Thammasat University, Pathum Thani, Thailand; Sirindhorn College of Public Health, Chonburi Province, Praboromarajchanok for Health, Workforce Development, Thailand; Department of Community Medicine and Family Medicine, Faculty of Medicine, Thammasat University. Pathum-Thani, Thailand

**Keywords:** aging, inappropriate prescription, hospitalization, adverse drug event

## Abstract

**Purpose:** To examine the association between potentially inappropriate medications (PIMs) use and the hospitalization rate in elderly Thai patients.

**Methods:** In this retrospective cohort study, we collected the electronic medical data of elderly patients aged 60 years and older who visited the outpatient department (OPD) at Thammasat University Hospital in Thailand in 2015. The patients were categorized into PIM and non-PIM users according to the Beers 2019 criteria. We calculated descriptive statistics for demographic variables. We also examined the association between PIM use and various different factors with hospitalization rate during follow-up using log-binomial regression. We calculated the relative risk for association between PIM use and other factors with the hospitalization rate.

**Results:** We collected data for a total of 32,261 patients. The majority of participants were female (59.65%) and had a mean age of 70.21 years (SD=7.88). Overall, 63.98% of the patients (n=20,641) were PIM users and 49.45% (n=15,952) received polypharmacy (≥5 medications). The most common PIM prescription was proton-pump inhibitors, which were 27.51% of all medications prescribed. We found that PIM use increased the risk of hospitalization by 1.31 times (adjusted RR=1.31, 95% CI: 1.21 – 1.41, p-value < 0.001). Other factors associated with a higher rate of hospitalizations included older age, male gender, polypharmacy, and a higher number of OPD visits.

**Conclusion:** PIMs were commonly prescribed to the elderly in the OPD, and were significantly associated with subsequent hospitalization. The provision of an alternative drug list can help physicians avoid prescribing PIMs to the elderly. If PIMs prescription is unavoidable, physicians should closely monitor patients for drug-related problems and deprescribe PIMs when they are no longer indicated.

## 1. Introduction

Thailand has become an aging society since 2002 [1]. This situation is of particular concern because the incidence of disease in the elderly is four times higher when compared to other age groups. Thus, the elderly are more prone to receiving potentially inappropriate medications (PIMs). PIMs are associated with drug-related problems (DRPs), such as adverse drug reactions (ADRs). Compared to younger people, the elderly are more vulneralbe to ADRs because of biological and physiological changes that accompany aging. PIMs in the elderly can lead to negative health outcomes in the long run and decrease their quality of life. [2-3]

The use of inappropriate medications in the elderly is associated with negative health outcomes, including increased hospitalization, adverse drug events, and higher resource utilization. A study in Japan found that 8% of patients with PIMs experienced adverse drug event. This study showed that benzodiazepines, which cause lightheadedness, somnolence, and sleepiness, were associated with increased risk of falls and fractures in the elderly [4]. A study in the Southeast region of the United States showed that the prevalence of DRPs was significantly higher among those who received PIMs compared than their counterparts not receiving PIMs. DRPs included alteration of consciousness, syncope, sleep disturbances, urinary incontinence, bradycardia, fall, fracture, dementia, hypotension, and gastritis. Many of these problems are associated with increased hospitalization risk [3].

The two commonly used criteria for defining PIMs are the American Geriatric Society Beers Criteria (AGS Beers Criteria) and STOPP/START (Screening Tool of Older person’s Prescriptions/Screening Tools to Alert doctors to Right Treatment. The AGS Beers Criteria for screening Potentially Inappropriate Medication were developed in the USA in 1991 to decrease inappropriate prescribing and adverse drug events in nursing homes. These criteria were recently updated in 2019. The Beers criteria were designed for elderly people aged 65 years and older. These criteria are classified by drug-related problems.[5]. The STOPP/START criteria for potentially inappropriate prescribing in older people was developed in Europe in 2008 (Version 1). The STOPP/START criteria were updated in 2014 (Version 2). The new version of this criteria included 80 STOPP criteria which were classified by the physiological system.[6]

The prevalence of PIMs using the Beers 2019 criteria in tertiary hospitals in China and India was 55.0% and 61.9%, respectively [7, 8]. The most common prescribed PIM according to Beers 2019 criteria in hospitalized patients in Brazil was metoclopramide, follow by omeprazole and regular insulin [9]. Similarly, the most prescribed PIMs according to the Beers 2019 criteria in elderly hospitalized patients in tertiary care were pantoprazole, omeprazole. and insulin [8].

Previous studies have reported an association between PIM use and hospitalization rates, ranging from 16%-90% increases in risk of hospitalization [10, 11]. To our knowledge, there are no previous studies regarding the association between PIM use and the hospitalization rate in Thailand. The purpose of this study was to examine the association between PIM use and the hospitalization rate in elderly Thai patients. In addition, we sought to identify additional factors associated with increased hospitalization rate.

## 2. Methods

### 2.1. Study design and participants

This retrospective cohort study examined the association between PIM use and the hospitalization rate in elderly Thai patients at Thammasat University Hospital, Thailand. In addition, we identified other factors associated with the hospitalization rate. All data were collected from electronic medical records between 2015 and 2017. Participants were elderly patients aged 60 years old and above who visited the outpatient department (OPD) at Thammasat University Hospital between January 2015 and December 2015. The age cutoff of 60 years old was chosen according to the definition of elderly in The Act on the Elderly, B.E. 2546 [1]. Patients were excluded if they were not diagnosed with any primary diagnosis, or did not receive any medications. We categorized the participants into PIM users and non-PIM users.

### 2.2. Data collection and variables

We collected patient data from electronic medical records at Thammasat University Hospital in 2015. We gathered data about the following factors based on our literature review: demographic data including age [12, 13], gender (male, female) [13, 14], marital status [13], and health insurance schemes [13]. We categorized age as 60-74 and ≥ 75 years based on a previous study [15]. The categories of marital status consisted of married, single, widow, divorced, and others. The categories for health insurance schemes consisted of civil servant medical benefit scheme, universal coverage scheme, cash, social security scheme, and others. The health insurance schemes dictated a patient’s access to a range of medications and services [16, 17].

We classified patient diagnoses according to the 10th Revision of International Classification of Disease and health-related problems (ICD-10) [18]. We collected data on the drugs prescribed to OPD patients. Data on the prescribed drugs consisted of name, dosage, drug administration, number of medication, and prescription date [5,8,19]. These data were used to define patients as either PIM users or non-PIM users. Prescriptions with five or more medications were defined as polypharmacy [19]. We classified the number of OPD visits as: 1-3 visits and ≥ 4 visits [20].

We collected hospitalization data from 2016-2017 for elderly patients. Hospitalization was defined as having at least one admission into any of the hospital wards, excluding the emergency department and ambulatory surgery, regardless of the length of stay, the number of admissions, and the nature of admission (elective or emergency).

### 2.3. PIMs

We used the AGS Beers 2019 criteria to define PIMs [20]. AGS Beers 2019 criteria consists of 5 categories: 1) PIM use in older adults, 2) PIMs use in older adults due to drug-disease or drug-syndrome interactions, 3) Drugs to Be Used with caution in older adults, 4) Potentially clinically important drug-drug interactions that should be avoided in older adults, and 5) Medications that should be avoided or have their dosage reduced with varying levels of kidney function in older adults [5]. PIMs in this study were identified by applying the two categories: 1) medication use in older adults and 2) medication to be used with caution in older adults. PIM users were defined as receiving at least one PIM in 2015.

### 2.4. Data analysis

Both descriptive and analytical analyses were performed. We calculated the mean and standard deviation (SD) for age, number of primary diagnoses, number of medications, and number of OPD visits. We calculated the proportion for different categories of gender, marital status, health insurance scheme, age group, group of the number of OPD visits, top five common diagnoses at the OPD, top five common prescribed PIMs, and the number of polypharmacy users. We used log-binomial regression to calculate the adjusted relative risk for the association between PIM use, gender, age, polypharmacy, and the number of OPD visits with hospitalization rate. Relative risk was suitable for cohort studies with common outcome [21]. We used a p-value of 0.05 to indicate statistical significance in this study. All analyses were conducted using STATA software version 14.2 (Stata, College Station, TX, USA).

## 3. Results

## 3.1. Participant Eligibility

A total of 35,051 older patients were eligible for this study between January to December 2015. Of these, 32,261 patients were included in the study. These patients were then categorized into PIM users and non-PIM users. We examined 2-years of follow-up hospitalization data (Fig 1).

**Fig. 1.**
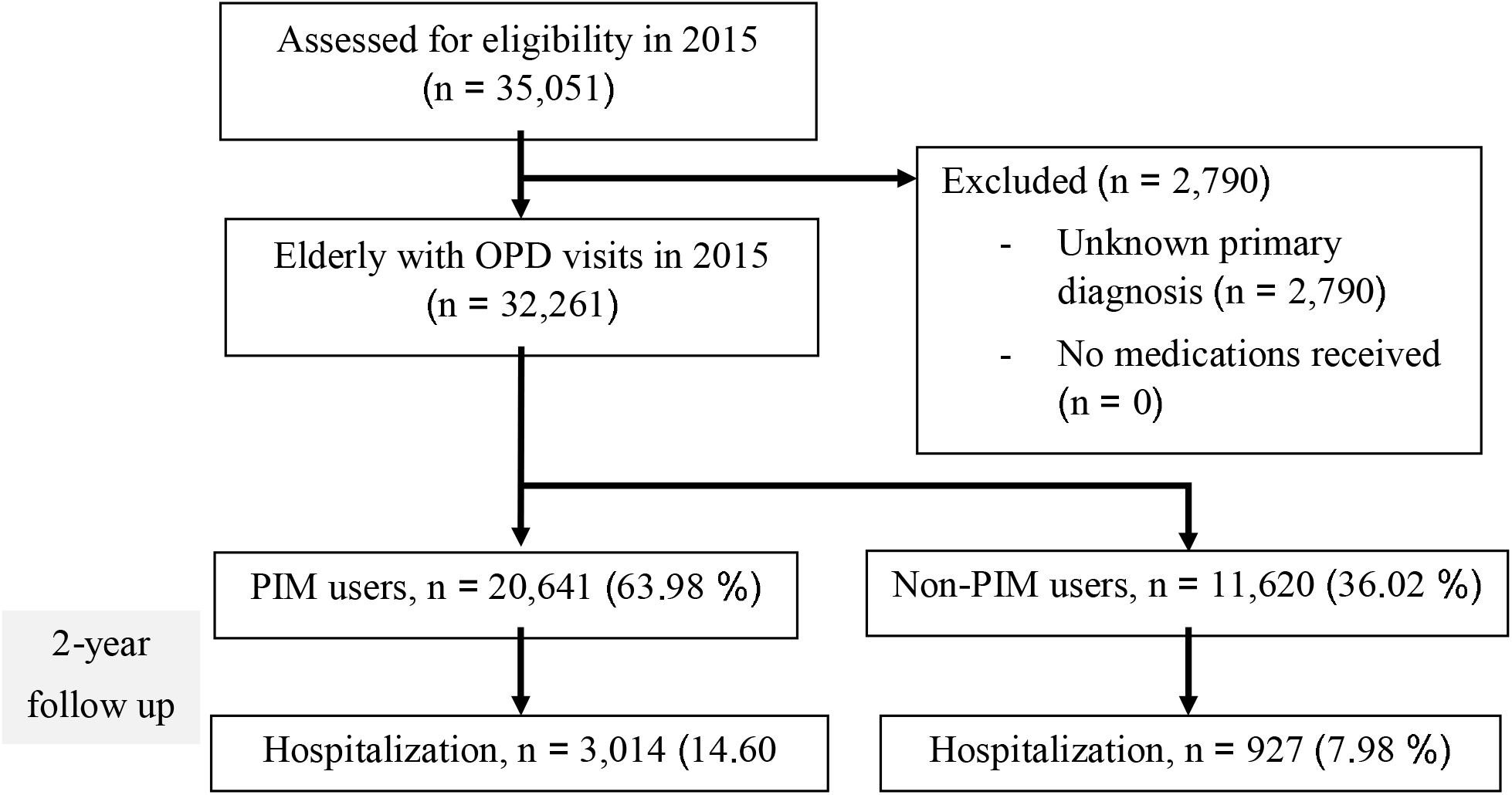
Retrospective cohort study design and study flow chart. Abbreviations: OPD = Out-patient department; PIMs = Potentially inappropriate medications by American Geriatric Socieity (AGS) Beers 2019 criteria

### 3.2. Patients’ characteristics

Of the 32,261 elderly patients included in our study, 19,244 (59.65 %) were female and 22,947 (71.13 %) were 60-74 years old. Most patients were married (70.05 %), used the civil servant medical benefit scheme (48.56 %), and were prescribed from one to four different medications (50.55 %) (Table 1). The mean number of the diagnoses at the OPD was two diagnoses per patient (1.96 ± 1.04). The most common primary diagnosis at OPD visits of both PIM and non-PIM users was essential (primary) hypertension (11.10 % and 11.02 %, respectively), followed by type 2 diabetes mellitus without complications (3.75 % and 4.38 %, respectively). Osteoarthritis was one of the top five diagnoses among PIM users but not among non-PIM users (Table 2).

**Table 1.**
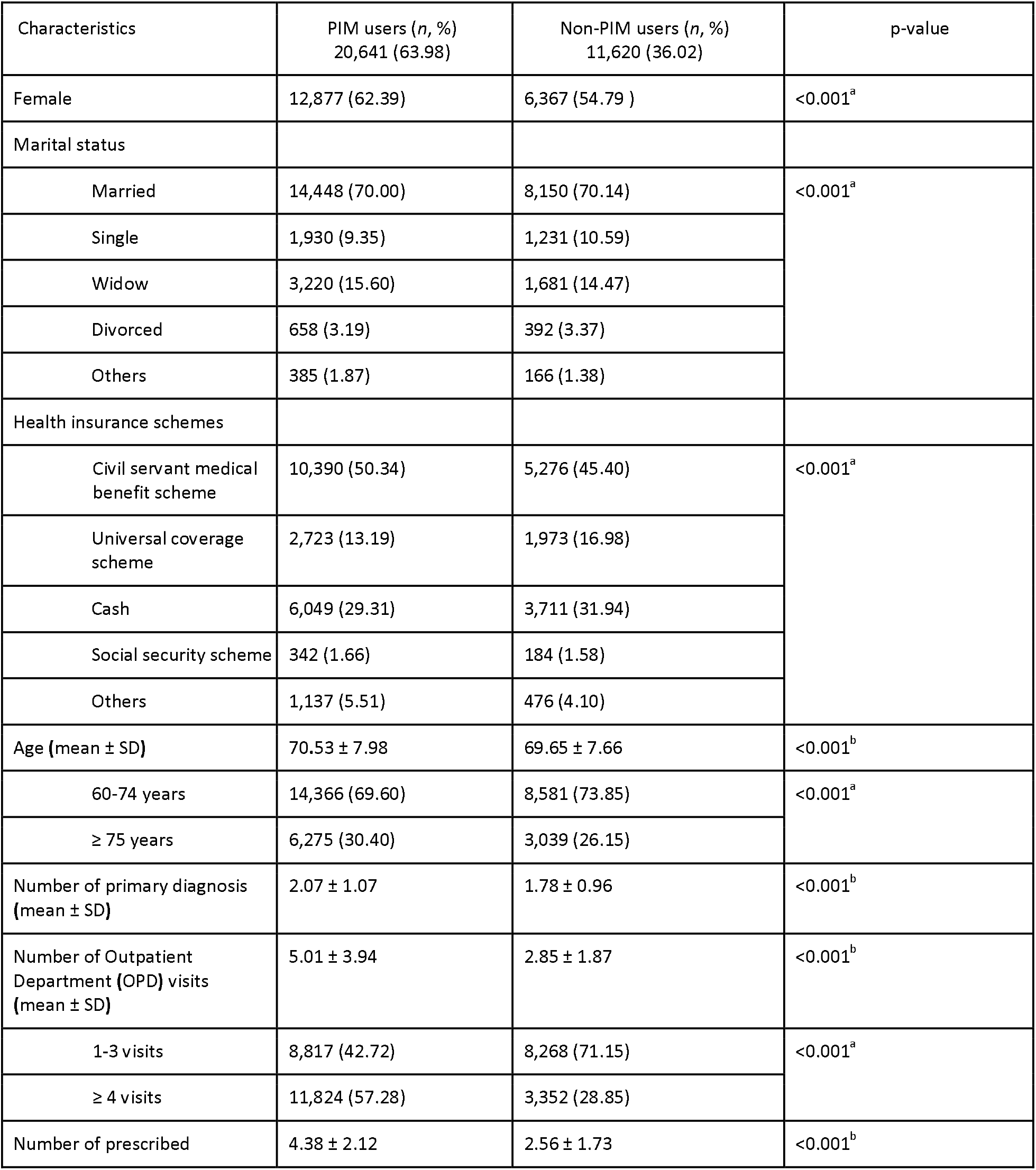
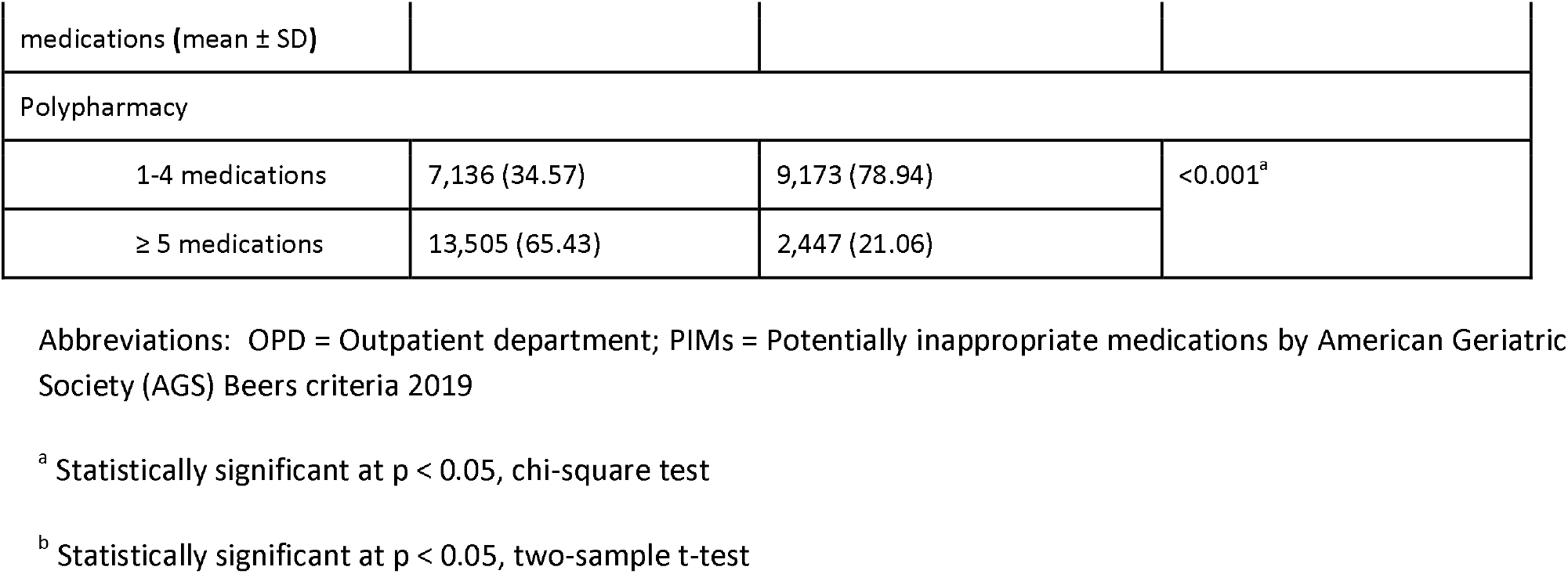
Descriptive statistics for study participants at Thammasat University Hospital, Thailand from 2015-2017 (*n*=32,261)

| Characteristics | PIM users (*n*, %)<br>20,641 (63.98) | Non-PIM users (*n*, %)<br>11,620 (36.02) | p-value |
| --- | --- | --- | --- |
| Female | 12,877 (62.39) | 6,367 (54.79) | <0.001<sup>a</sup> |
| Marital status |  |  |  |
| Married | 14,448 (70.00) | 8,150 (70.14) | <0.001<sup>a</sup> |
| Single | 1,930 (9.35) | 1,231 (10.59) |  |
| Widow | 3,220 (15.60) | 1,681 (14.47) |  |
| Divorced | 658 (3.19) | 392 (3.37) |  |
| Others | 385 (1.87) | 166 (1.38) |  |
| Health insurance schemes |  |  |  |
| Civil servant medical benefit scheme | 10,390 (50.34) | 5,276 (45.40) | <0.001<sup>a</sup> |
| Universal coverage scheme | 2,723 (13.19) | 1,973 (16.98) |  |
| Cash | 6,049 (29.31) | 3,711 (31.94) |  |
| Social security scheme | 342 (1.66) | 184 (1.58) |  |
| Others | 1,137 (5.51) | 476 (4.10) |  |
| Age (mean ± SD) | 70.53 ± 7.98 | 69.65 ± 7.66 | <0.001<sup>b</sup> |
| 60-74 years | 14,366 (69.60) | 8,581 (73.85) | <0.001<sup>a</sup> |
| ≥ 75 years | 6,275 (30.40) | 3,039 (26.15) | |
| Number of primary diagnosis (mean ± SD) | 2.07 ± 1.07 | 1.78 ± 0.96 | <0.001<sup>b</sup> |
| Number of Outpatient Department (OPD) visits (mean ± SD) | 5.01 ± 3.94 | 2.85 ± 1.87 | <0.001<sup>b</sup> |
| 1-3 visits | 8,817 (42.72) | 8,268 (71.15) | <0.001<sup>a</sup> |
| ≥ 4 visits | 11,824 (57.28) | 3,352 (28.85) | |
| Number of prescribed | 4.38 ± 2.12 | 2.56 ± 1.73 | <0.001<sup>b</sup> |
| medications (mean ± SD) |  |  |  |
| Polypharmacy |  |  |  |
| 1-4 medications | 7,136 (34.57) | 9,173 (78.94) | <0.001<sup>a</sup> |
| ≥ 5 medications | 13,505 (65.43) | 2,447 (21.06) |  |

Abbreviations: OPD = Outpatient department; PIMs = Potentially inappropriate medications by American Geriatric Society (AGS) Beers criteria 2019
<sup>a</sup> Statistically significant at $p < 0.05$ , chi-square test
<sup>b</sup> Statistically significant at $p < 0.05$ , two-sample t-test

**Table 2.**
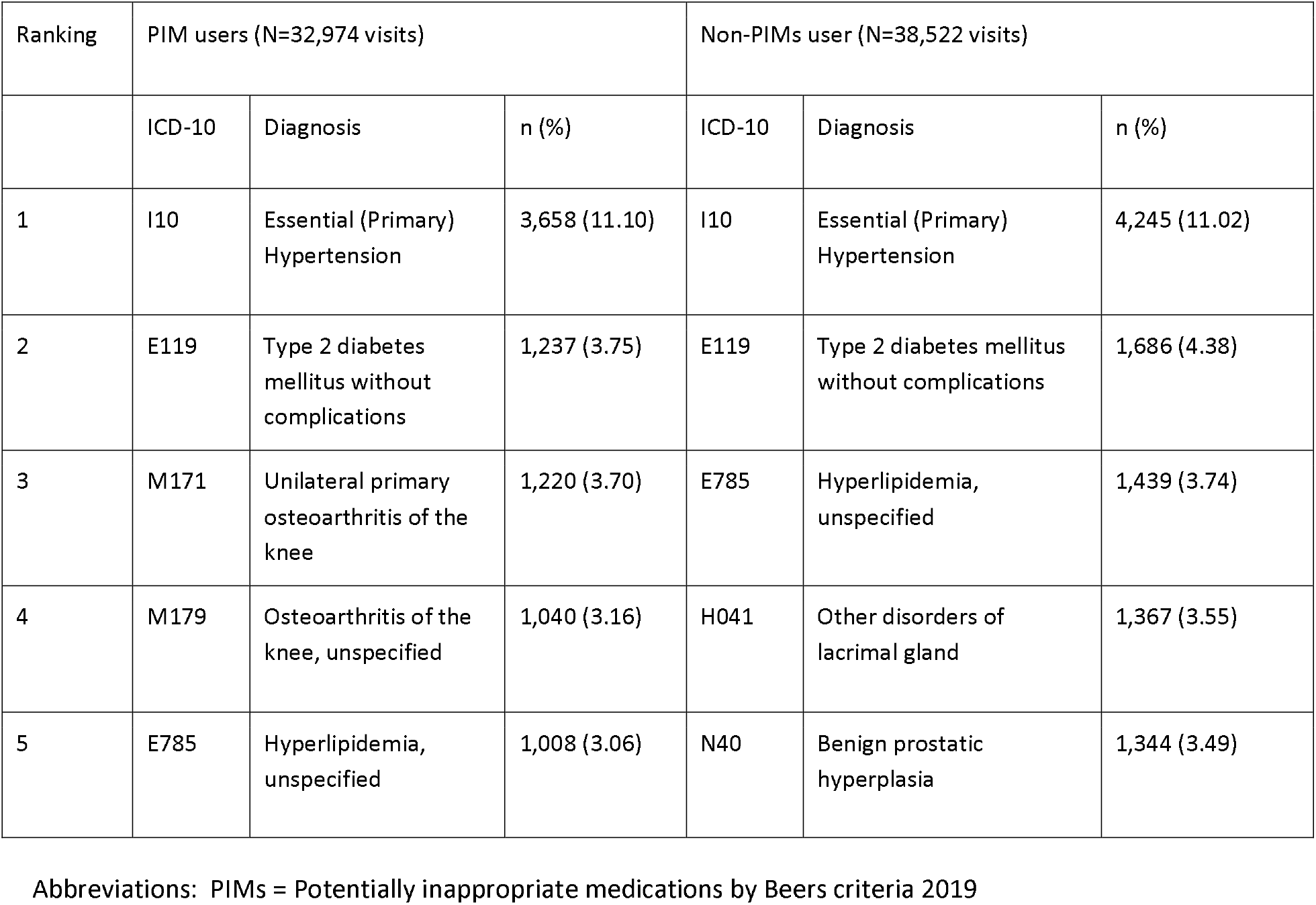
Top primary diagnoses of PIM and non-PIM users at the Outpatient Department (OPD) in 2015 at Thammasat University Hospital, Thailand.

### 3.3. Prevalence of Potentially Inappropriate Medications

Nearly 64% of all study participants had one or more PIMs prescribed according to the AGS Beers 2019 criteria. On average, PIM users were prescribed 4.38 medications (SD=2.12) for each OPD visit. Compared to non-PIM users, PIM users visited OPD more frequently (5.01 ± 3.90 visits versus 2.85 ± 2.12 visits in non-PIMs users, p < 0.001). Of the PIM users, 13,505 (84.66 %) patients were prescribed with polypharmacy (≥ 5 medications) compared to 2,447 (15.34 %) patients in non-PIM users (p < 0.001) (Table 1).

The top three most frequently prescribed PIMs in elderly adults were proton-pump inhibitors (PPIs) (29,023 prescriptions, 27.51 %), followed by non-steroidal anti-inflammatory drugs (NSAIDs) (9,119 prescriptions, 8.64 %), and short- and intermediate-acting benzodiazepines (9,063 prescriptions, 8.59 %). In addition, the top three drugs to be used with caution in older adults were diuretics (11,040 prescriptions, 10.46%), tramadol (8,706 prescriptions, 8.25%), and tricyclic antidepressants (4,129 visits, 3.91%) (Table 3).

**Table 3.** Top potentially inappropriate medications (PIMs) and drugs to be used with caution in older adults prescribed at Outpatient Department (OPD) in 2015 at Thammasat University Hospital, Thailand. (_n_=105,509 prescriptions)

| PIM use in older adults | *n* (%) |
| --- | --- |
| Proton-pump inhibitors (PPIs) | 29,023 (27.51) |
| Non-steroidal anti-inflammatory drug (NSAIDs),<br>cyclooxygenase non-selective | 9,119 (8.64) |
| Short and intermediate-acting Benzodiazepines | 9,063 (8.59) |
| Skeletal muscle relaxants | 8,881 (8.42) |
| Anticholinergics | 6,746 (6.39) |
| **Drugs to Be Used with caution in older adults** |  |
| Diuretics | 11,040 (10.46) |
| Tramadol | 8,706 (8.25) |
| Tricyclic antidepressants | 4,129 (3.91) |
| Selective serotonin reuptake inhibitor | 3,506 (3.32) |
| Antipsychotics | 2,299 (2.18) |

Abbreviations: PIMs = Potentially inappropriate medications by Beers criteria 2019

### 3.4. The association between PIMs and hospitalizations

PIM users had a higher hospitalization rate compared with non-PIMs users (3,014 patients (14.60 %) and 927 patients (7.98 %), respectively). PIM use was associated with a 1.31-fold increase in hospitalization rate (adjusted RR=1.31 [95 % CI 1.21–1.41]). Other factors also associated with hospitalization rate, included being 75 years old or older (adjusted RR 1.49 [95 % CI 1.41 – 1.59]), male gender (adjusted RR 1.12 [95 % CI 1.05 – 1.18]), polypharmacy (adjusted RR 1.48 [95 % CI 1.37 – 1.59]), and having 4 or more OPD visits (adjusted RR = 1.68 [95 % CI 1.57 – 1.79]) (Table 4).

**Table 4.** Crude and adjusted relative risks of hospitalization for age, gender, polypharmacy, PIMs prescription, and number of OPD visits in 2016-2017 at Thammasat University Hospital, Thailand.

| Variables | Crude relative risk (95% CI) | Adjusted relative risk (95% CI) |
| --- | --- | --- |
| Age |  |  |
| <75 | 1 | 1 |
| ≥ 75 years | 1.66 (1.56-1.76) | 1.49 (1.41 – 1.59) |
| Gender |  |  |
| Female | 1 | 1 |
| Male | 1.10 (1.03-1.16) | 1.12 (1.05 – 1.18) |
| Polypharmacy |  |  |
| No | 1 | 1 |
| Yes | 2.09 (1.96-2.23) | 1.48 (1.37 – 1.59) |
| PIMs prescription |  |  |
| No | 1 | 1 |
| Yes | 1.83 (1.71-1.96) | 1.31 (1.21 – 1.41) |
| Number of OPD visits |  |  |
| 1-3 | 1 | 1 |
| ≥ 4 | 2.14 (2.01-2.27) | 1.68 (1.57 – 1.79) |

Abbreviations: OPD = Out-patient department; PIMs = Potentially inappropriate medications by Beers criteria 2019

## 4. Discussion

In our retrospective cohort study, we found that the prevalence of PIM was high. We also observed that PIM prescription in the elderly was associated with increased risk of hospitalization. The most commonly prescribed PIMs in older adults were PPIs. The most frequently prescribed PIMs for drugs to be used with caution in older adults were diuretics. Other factors associated with increased hospitalization were older age, male gender, having four or more OPD visits during the 2-year follow-up period, and polypharmacy.

Regardless of the criteria used to define PIMs and the patient setting, the prevalence of PIM prescription in our study was congruent with other studies. Investigators found the prevalence of PIMs was 56.88% at a Jordanian elderly outpatient at a tertiary hospital, according to the Beers 2015 criteria [22]. Also using the AGS Beers 2015 criteria, another study found the prevalence of PIMs was 57.7% in an ambulatory care setting in a large tertiary hospital in Saudi Arabia [23]. Using the AGS Beers 2019 criteria, Wang et al. found that 55.0% of hospitalized elderly patients in a tertiary hospital had PIM upon hospital admission [7]. Also, using AGS Beers 2019 criteria, Sharma et al. found the prevalence of PIM was 61.9% among older adults admitted to a tertiary care postgraduate teaching hospital in India [8]. The slight variation in the prevlanece of PIMs in different studies may be due to differences in the Beers criteria being used, the characteristics of the patients, the disease status, the hospital settings, the difference in the prescribing patterns, and the availability of the drugs marketed in different countries [24].

To the best of our knowledge, no previous study has been conducted using the AGS Beers 2019 criteria in Thailand. However, the prevalence of PIMs prescription in the present study was relatively high compared to other studies in Thailand. For example, the prevalence of PIMs was 40.4% in a study in the primary care unit of a university hospital in Southern Thailand [20] and 59.0% in a semiurban primary care unit [25]. Compared to the previous studies, the patients in our study had more complex diseases, which put them at risk of being prescribed PIMs and polypharmacy. Furthermore, most of the patients in our study used the civil servant medical benefit scheme, which gave them access to a larger variety of medications [16, 17]. In contrast, in other studies in Thailand, most of the patients used the universal health coverage scheme, which limits the types of medications that patients can receive [20, 25].

PPIs were the most commonly prescribed PIM in our study. Similarly, researchers found that 39.4% of all PIM prescriptions at family and community medicine clinics in Saudi Arabia were for PPIs [26]. The Beers 2019 criteria recommend avoiding PPIs for more than 8 weeks, except in high-risk patients or patients with a clear indication such as erosive esophagitis or pathological hypersecretory condition. Long-term use of PPIs increases the susceptibility of Clostridium Difficile infection, bone loss, and fractures [5]. In our study, the high prevalence of PPI use was consistent with the higher prevalence of osteoarthritis (OA) of the knee (ICD M171, M179) among PIMs users compared to non-PIMs users. Concomitant use of PPIs with cyclooxygenase (COX) non-selective NSAIDs in older patients is recommended to prevent gastrointestinal bleeding. In addition, COX-2 selective NSAIDs without PPIs are a better alternative to this age group [27]. In Thailand, omeprazole enteric-coated caps were listed in the national list of essential medicine, so it was widely available to everyone free of charge [28]. Long-term omeprazole is also indicated in patients with chronic dyspepsia and gastrointestinal reflux disease [29].

The most commonly prescribed medication to be used with caution in our study was diuretics. This finding was consistent with the high prevalence of essential hypertension and chronic kidney disease in our study, as diuretics were indicated in treating both diseases [30, 31]. Similar to our study, researchers found that diuretics were the most commonly prescribed PIM for patients at the family and community medicine clinics in Saudi Arbia (25.2% were prescribed diuretics) [26]. The AGS Beers 2019 criteria state that diuretics increase the risk of developing the syndrome of inappropriate antidiuretic hormone and hypernatremia. Clinicians should closely monitor a patient’s serum sodium level when starting diuretics or changing the medication dosage [5].

We found that PIM use is associated with increased the rate of hospitalization. Previous studies’ reports on this association were inconclusive. Studies in Taiwan [19], South Korea [32], and Switzerland [33] found that PIM use, based on the AGS Beers 2012 criteria, was associated with increased hospitalization rate (27% in Taiwan, 25% in South Korea and 13% in Switzerland). In contrast, studies in Japan and Italy did not find that PIM use was associated with greater hospitalization [34, 35]. The variation between these studies’ findings may be due to the lack of investigation on the causes of hospital admission, such as the patient’s pre-existing medical conditions, the exacerbation of the underlying disease, and the adverse drug events of PIMs. To the best of our knowledge, no other previous studies have investigated the association of PIMs with hospitalization by using the AGS Beers 2019 criteria. Therefore, we cannot compare our study results with other studies..

PIM use increases the risk of adverse drug events, and thus increases the rate of hospitalization. Previous research in Japan reveals that the prevalence of PIM-induced adverse drug events was 8%. Benzodiazepines, which cause dizziness, somnolence, and sleepiness, can increase the risk of falling in older adults [4]. Also, PIM use is associated with increased prevalence of drug-related problems relative to non-PIM use, such as alteration of consciousness, syncope, sleep disturbance, fall, gastrointestinal bleeding, and gastritis [3]. These conditions increase the risk of hospitalization.

The results of the log-binomial regression in our study showed that the hospitalization rate was associated with older age, being male, polypharmacy, and having 4 or more OPD visits within a 2-year follow-up period. Older age is associated with adverse outcomes of the patient’s pre-existing conditions and diseases [24]. The hazard ratio hazard rate for increased age was 1.02 in a study in Sweden [36]. Similarly, Alexnder et al. in the United States found that a ten-year increase in age was associated with a 1.16-fold increase in risk of hospitalization [12].

Interestingly, our study found that male patients had a higher risk of hospitalization than female patients. This occurred despite the fact that females had a higher PIM prevalence. Based on our findings, we suspect that being male is a factor which increases hospitalization rate, regardless of PIMs. Further investigation of this phenomenon should be conducted.

The association between the number of OPD visits and hospitalization in our study was consistent with a previous study. This study documented that over six OPD visits increased risk of hospitalization by 1.6 times [13]. Patients with frequent OPD visits usually have polypharmacy, PIMs, and duplicated medications [37]. The nature of their health conditions is generally severe, chronic, or complicated. This situation makes patients with frequent OPD visits particularly prone to hospitalization.

We found a high prevalence of polypharmacy among our study participants. We also confirmed that polypharmacy was associated with increased risk of hospitalization. Other previous studies reported the high prevalence of polypharmacy in Thailand [38] and Kuwait [39]. Patients with polypharmacy are at risk of hospitalization because they usually have multiple comorbidities and one or more PIMs [40]. Using the National Health Insurance Research Database, a study in Taiwan found that receiving 5 to 9 medications and more than ten medications increased the hospitalization rate by 1.34 times and 1.98 times, respectively [19]. Another study in Japan found that elderly patients with polypharmacy were more likely to be hospitalized than those without polypharmacy (OR 2.11, 95% CI: 1.03-4.43) [41].

Our study has some limitations. First, we did not have data on the patients’ overall drug use outside of medical records from Thammasat University Hospital. Consequently, it is possible that we underestimated the prevalence of PIM. Secondly, we did not take into consideration the cause of hospitalization. Thus, the association between PIMs use with hospitalization could be overestimated. Lastly, the prevalence of PIMs could differ if we defined PIMs according to all the categories in the AGS Beers 2019 criteria.

## 5. Conclusion

PIMs are prescribed to 63.98% of the elderly in the OPD. Among our study participants, PIMs were significantly associated with subsequent hospitalizations, with the adjusted relative risk of 1.31 (95%CI 1.21 – 1.41, p-value < 0.001). When prescribing medications, physicians should thoroughly review a patients’ medication record and prescribe PIMs after evaluating potential risk and benefits of PIMs. A provision of an alternative drug list can help physicians avoid prescribing PIMs to the elderly. Most importantly, physicians should be cautious about prescribing PIMs. They should be aware of the drug-related problems due to PIMs in order to prevent and detect these problems.

Future studies should be conducted to explore the causes of hospital admission, length of stay, the economic burden of disease, and hospitalization outcomes in PIM users relative to non-PIM users. Comparing the treatment outcome and efficacy of PIM and non-PIM in the elderly patients should also be investigated..

## Data Availability

Available upon request

## Declaration

### Availability of data and material

Available upon request

### Ethics approval

The Ethical Review Sub-Committee Board for Human Research of Faculty of Medicine, Thammasat University approved this study (MTU-EC-CF-0-221/61).

### Consent to participate

Not applicable

### Consent for publication

Not applicable

### Authors’ Contribution

Vorawee Varavithya: study design, data collection, data analysis, manuscript writing

Chayanee Tirapat: study design, data collection, data analysis, manuscript writing

Penpitcha Rojpibulstit: study design, data collection, data analysis, manuscript writing

Panadda Poovichayasumlit: study design, data collection, data analysis, manuscript writing

Vanida Prasert: study design, data analysis, manuscript writing

Pasitpon Vatcharavongvan: study design, data analysis, manuscript writing

